# Impact of a Clinical Orientation Programme as perceived by new entrant international medical graduate resident doctors partaking in the programme: Results from an NHS district hospital survey

**DOI:** 10.1101/2024.10.29.24316389

**Authors:** Syed Rehan Quadery, Thomas John, Simon Winn

## Abstract

**Background:** International Medical Graduates (IMGs) constitute 40% of the NHS resident doctor workforce. However, there is no documented clinical orientation programme (COP) facilitating their transition into the workplace. We developed and implemented a structured COP to support new entrant IMGs.

**Method:** Survey data was collected from new entrant IMGs in medicine who completed the COP between December 2017 and December 2019. In addition, formal feedback was collected from supervising registrars during the COP period. Data was analysed, and a framework was developed to assess impact using realist evaluation.

**Results:** 97% (31of 32) of IMGs completed the survey. Results revealed 84% were satisfied with the programme, 81% felt confident they were prepared to begin clinical posts at completion of the programme, and 90% would recommend this programme. In addition, the overall comments from the IMGs were positive and supportive of the COP. Moreover, feedback from supervising registrars was complementary. It was seen that 87% of IMGs remained Trust employees with a mean tenure of 13 months. Of those eligible, 90% completed their annual appraisal.

**Conclusion:** This report confirms that Trust in this COP results in a high degree of satisfaction, workforce retention and performance of new entrant IMGs. Larger scale implementation is recommended.

## Background

The medical workforce of the National Health Service (NHS) consists of 63% locally trained and 37% overseas doctors, of whom 11% originate from the European Union (EU) and 26% are International Medical Graduates (IMGs).^1, 2^ IMGs are doctors who have completed their primary medical qualifications from countries outside the UK and the EU.^1, 2^ Current estimates point to a significant shortage in the NHS medical workforce which might lead to compromises in patient care.^3^ It has also been speculated that this situation might be exacerbated due to the reduced migration of EU doctors to the United Kingdom following Brexit.^4^ To combat this eventuality, the interim NHS People Plan proposed increasing the number of locally trained doctors as a measure to bolster the existing medical workforce. However, there will likely be a delay of several years before these changes demonstrate an effect.^5^ Therefore, recruitment and retention of IMGs is an attractive short to medium term solution to fill staffing gaps and mitigate the workforce shortfall to help maintain patient care.^5^

Once recruited, IMGs in their first clinical role within the NHS face multiple challenges. These include but are not limited to factors such as adjusting to a new culture, training systems, language barriers, medico-legal framework, professional duties, and skills.^6^ These challenges have been shown to impact clinical performance, career progression, personal wellbeing, and patient care.^7^

Furthermore, new entrant IMGs might not be considered ready to perform out of hours clinical duties because of limited formal assessment of their competence and knowledge of local systems. In this context, a meticulously designed Clinical Orientation Programme (COP) can be utilized as a valuable tool anticipate and mitigate such challenges and facilitate integration of the IMGs into the NHS. However, there is no evidence of a nationally accredited programme available for new entrant IMGs working in General Internal Medicine (GIM) in the NHS.^2^ Also, a thorough search of the existing literature reveals no published data available assessing the impact of such a COP on this staff group. We have recently described a quality improvement intervention of recruitment and retention of resident doctors with a focus on a structured COP for new entrant IMGs in the department of GIM at two district general hospitals.^8^ Understanding the perceptions of IMGs regarding this process ensures continued quality improvement and facilitates early integration into the workplace which is fundamental to ensuring a stable resident doctor workforce. This report therefore aims to describe the perceptions of IMGs undergoing such a COP.

## Methods

### Study participants

Between 1st December 2017 and 15th December 2019, 33 new entrant IMGs participated in the COP. Of note, 49% (n=16) participants completed the Trust’s honorary clinical attachment programme before starting the COP. Of the IMGs, 97% (n = 32) successfully completed the COP and were invited to complete the survey. The COP targets are shown in (Table 1).

**Table 1:** Targets in the clinical orientation programme.

| SN | Competency targets |
| --- | --- |
| 1 | Using medical imaging technology such as picture archiving and communication system (PACS) for reviewing and commenting on radiological results |
| 2 | Using online software via trust intranet such as clinical manager (iCM) for electronic requesting, results reporting, clinical documents for creating discharge summaries, TTO prescribing and electronic in- patient prescribing of medications and online speciality referrals |
| 3 | To confirm drug doses and ensure good antibiotic stewardship using intranet resources including British National Formulary and Trust antibiotic guidelines |
| 4 | Using a software system such as VitalPAC for assessing vital signs of patients by based on national early warning score system in recognizing deteriorating patients in hospital |
| 5 | Using nhs.net email for sending and receiving patient sensitive data within and outside the organization |
| 6 | Using an Arterial Blood Gas analyser including its access code |
| 7 | Answering and responding to bleeps |
| 8 | Effective ward round documentation including a checklist |
| 9 | Ability to make telephonic and face to face referrals to other specialities in and outside the organization including radiology and microbiology |
| 10 | To access the medical clerking proforma and clerking and presenting patients in the acute unselected medical take to consultants |
| 11 | Using software in the management and updating of the acute unselected take list via the Acute Medicine Unit Bed Manager programme |
| 11 | To manage acute medical emergencies * |
| 12 | Basic life support provider |
| 13 | Knowledge and implementation of DNAR, PTEP, end of life care and advanced care planning |
| 14 | Management of complex elderly patients with multiple co-morbidities |
| 15 | Performing of core procedures as described at the end of Foundation programme # |
| 16 | Awareness of Trust guidelines and protocols including VTE thromboprophylaxis, dementia screening, medication reconciliation, treatment escalation discussion and planning and accessing Trust clinical guidelines via Trust intranet |

Definition of abbreviations: TTO=to take out; DNAR=do not attempt resuscitation; PTEP=personalized treatment escalation plan; VTE=venous thromboembolism;
\* Acute medical emergencies include acute myocardial infarction, acute pulmonary embolism, pneumonia, acute severe asthma, acute exacerbation of COPD (including NIV), acute kidney injury and hyperkalaemia, upper GI bleed, comatose patient/low Glasgow coma scale, sepsis and septic shock, acute pulmonary oedema, headache with Acute subarachnoid haemorrhage / meningitis, status epilepticus and drug overdose (ie paracetamol).

### Data collection and analysis

A follow-up online survey via a Google Form was conducted to collect quantitative and qualitative data. The survey was open for 72 hours and closed on the 25th of December 2019. We used the realist evaluation approach for collecting and analysing both quantitative and qualitative data.^9^ Based on realist evaluation, the three concepts of Mechanisms (i.e., what makes a programme works?), Contexts (i.e., what are the conditions in which the mechanisms are operated?), and Outcomes (i.e., what are the observed outcomes of the programme?) were used to develop and test CMO Configurations (Where Contexts + Mechanism = Outcomes). In our survey, Contexts had one measure of current working conditions (3 items); Mechanisms had six measures of receiving training (1 item), co-worker support (1 item), management support (3 items), tailored programme fit for individual needs (1 item), participation (3 items), and assessment targets (1 item); And Outcomes had three measures of perception and awareness (9 items), work engagement (1 item), and wellbeing (3 items).^10^ Candidate responses were obtained on a five-point Likert scale with scores ranging from 1 (strongly agree) to 5 (strongly disagree). Descriptive data were presented using mean and standard deviation. Reliability of the survey was presented using Cronbach’s alpha.

Qualitative data was also collected under the final comments section as free text. Formal peer feedback was obtained from supervising middle grade doctors while on call in a supernumerary role. Feedback centred on clinical performance of individual IMGs and their ability to work more independently in their appropriate roles as part of the quality assurance process. To collect and analyse data, we followed a realist evaluation by using content analysis to extract themes of Contexts, Mechanisms, and Outcomes and then synthesise these themes to develop CMO Configurations^. 11^ Furthermore, demographics and programme related information was collected.

### Research Ethics

Ethical approval was waived by the Health Research Authority situated at 2 Redman Place, Stratford, London, E201JQ, bearing reference number 48/81 dated 28/05/2021. The HRA advised that a REC review would not be required for projects which only involve NHS staff (as opposed to patients) who are recruited into the study by virtue of their professional role. In addition, we used the decision tool “ does my study require REC review” to confirm that a REC review was not required.

## Results

A total of 31 of 32 IMGs (97%) responded to the survey. Of the participants, 45% (n=15) were female, 75% (n= 24) aged 26-35 years, 42% (n=14) married. We found that 80% (n=25) of IMGs were recruited at SHO grade, the remaining 20% (n=6) were registrar grade. It was seen that 87% (n=28) remain employees at the Trust with overall duration of tenure 13±8months. Of the eligible resident doctors, 90% completed their Annual Trust Appraisal. The survey revealed 84% (n=26) were satisfied with the COP, 81% (n=25) confident they were well-prepared to begin their posts at the end of the COP and 90% (n=28) would recommend this COP to friends and colleagues. Comprehensive results are shown in (Table 2). Overall survey reliability was found to be good (Cronbach’s alpha = 0.89). Qualitative feedback data from new entrant IMGs and peer feedback data from middle grade doctors supervising the IMGs during their COP are shown in (Table 3) and presented as themes (Context, Mechanism, Outcome), subthemes (e.g., senior mentoring as Mechanism) and comments in quotation. (Table 4) shows a range of CMO Configurations extracted from our data analysis. Achievements of new entrant IMGs during their employment with the Trust is shown in (Table 5).

**Table 2:** Quantitative feedback from new entrant International Medical Graduate.

| Parameters |  | Strongly Agree/<br>Agree %(n) |
| --- | --- | --- |
| **Context** |  |  |
| *Current working conditions* |  |  |
| 1 | You experienced bullying, harassment or intimidation during your placement. | 13 (4) |
| 2 | You had to leave late on a frequent and regular basis. | 26 (8) |
| 3 | The frequency and intensity of out of hours work during the clinical orientation period was manageable | 81 (25) |
| **Mechanisms** |  |  |
| *Receiving training* |  |  |
| 1 | You received training and access to the requisite Trust IT systems in a timely manner | 71 (22) |
| *Co-workers' support* |  |  |
| 1 | You received peer mentoring to help you achieve required goals throughout your placement. | 74 (23) |
| *Management support* |  |  |
| 1 | You received adequate clinical supervision (from consultants) during your placement. | 90 (28) |
| 2 | Your designated senior mentor provided you with guidance and maintained contact throughout as a means of consistent feedback. | 71 (22) |
| 3 | You received pastoral support throughout your placement. | 61 (19) |
| *Tailoring programme to fit individual needs* |  |  |
| 1 | Training arrangements were tailored to individual needs. | 65 (20) |
| *Participation* |  |  |
| 1 | There were opportunities to perform core procedures under supervision. | 84 (26) |
| 2 | There was sufficient opportunity to present patient cases to senior colleagues with feedback. | 90 (28) |
| 3 | Departmental teaching and in-house courses were made available within the Trust. | 77 (24) |
| *Clear Assessment Target* |  |  |
| 1 | The assessment targets were specific, measurable, achievable, relevant and time bound. | 74 (23) |
| **Outcome** |  |  |
| *Perceptions and awareness* |  |  |
| 1 | Prior to starting your programme clear information was provided. | 68 (21) |
| 2 | During your placement a holistic overview and general understanding of how the NHS functions (hospital and community) was provided. | 71 (22) |
| 3 | Since being placed with the Trust, you have a better understanding and overview of the Trusts systems, policies and processes (both clinical and non-clinical). | 87 (27) |
| 4 | During your placement you gained insight into the acute unselected medical take model and achieved the competencies for out of hours working. | 81 (25) |
| 5 | The duration of your orientation was appropriate to your learning needs. | 71 (22) |
| 6 | You were provided with learning in a supportive environment. | 81 (25) |
| 7 | You were provided with learning resources adequate to your training needs during the placement. | 84 (26) |
| 8 | You feel confident that you were adequately prepared to begin your post at the end of your orientation. | 81 (25) |
| 9 | Staff accommodation during your orientation reduced concerns regarding personal security, travel time and cost (between accommodation and work) and social isolation. | 42 (13) |
| *Work engagement* |  |  |
| 1 | You would recommend our programme to a friend or colleague. | 90 (28) |
| *Wellbeing* |  |  |
| 1 | You felt appreciated by the staff working in the organization during your placement. | 71 (22) |
| 2 | You were welcomed into the work area where you were based. | 77 (24) |
| 3 | You are satisfied with the programme. | 84 (26) |

**Table 3:** Qualitative feedback from new entrant international medical graduates (IMGs) and middle grade doctors supervising them during the clinical orientation period.

| **Feedback from new entrant IMGs about the clinical orientation programme (COP)** |  |  |  |
| --- | --- | --- | --- |
| **SN** | **Themes** | **Subthemes** | **Comments** |
| 1 | Mechanism | Senior mentoring | "Excellent mentoring program that would equip any new IMG to get integrated into the NHS system." |
| 2 | Mechanism | Pastoral care | The pastoral support was one of the key features of the programme." |
| 3 | Outcome | Awareness of contextual factors | "Orientation was very helpful in getting to know the system and I believe it should be mandatory for everyone joining first time in the NHS." |
| 4 | Context<br>Outcome | Lack of Peer mentoring<br>Self-efficacy | " Assigning a volunteer peer mentor who is at the same level to new IMGs during orientation period, would greatly build their confidence and achieve faster learning." |
| 5 | Context | Learning resources<br>(Lack of exposure to out of hours work) | "Could have been better in a few areas like improving my exposure to the acute take and ward on call rotas as supernumery. . I fed this back at the time and this was put in place for later doctors who did this programme." |
| 6 | Mechanism | Clinical supervision | "I must say my consultants were amazing and supportive and they helped me whenever I was stuck. Got to learn a lot from them as well." |
| 7 | Outcome<br>Outcome | Satisfaction<br>Recommending to friends and colleagues | "It is an excellent programme. I will highly recommend this orientation programme to my friends and will definitely help others in this programme to achieve their goals." |
| **Peer (registrar) feedback for IMGs regarding their performance during clinical orientation programme (COP)** |  |  |  |
| **SN** | **Themes** | **Subthemes** | **Comments** |
| 1 | Outcome | Quality assurance | "Dr X has demonstrated reasonable skills in managing the take and on-calls." |
| 2 | Outcome | Quality assurance | "I did an on call with Dr X. He was able to prioritise the take and handled everything in a smooth and an efficient way. " |
| 3 | Outcome | Quality assurance | "Well run take with appropriate management of patients. " |
| 4 | Outcome | Quality assurance | "He is performing at the stage of an early ST3 registrar." |
| 5 | Outcome | Quality assurance | "Dr X was very competent during my weekend on call as a supplementary SHO, where he performed his jobs independently and knew when to escalate, but overall was able to perform the job very well." |
| 6 | Outcome | Quality assurance | "I found that he's confident, has good clinical knowledge to make key decisions and remains calm and composed even when under stress. All of these I feel are essential to be a medical registrar and I feel Dr X will be a good and efficient medical registrar." |
| 7 | Outcome | Quality assurance | " I thought he did a very good job, he remained calm and objective throughout despite a flurry of referrals at the end of the day. He also took an individual interest in the team's welfare ensuring everyone had both eaten and had a break. He clearly is a clinician of many years' experience and his medical knowledge is sound. " |

**Table 4:** Context-mechanism-outcome Configuration.

| Contexts | Mechanism | Outcomes |
| --- | --- | --- |
| Shortage of JDs resulting in increased recruitment of temporary staff and inconsistent clinical care | Employing new entrant IMGs and providing a clinical orientation programme | Successful orientation of new entrant IMGs resulting in their improved clinical performance and satisfaction at an early stage resulting in better career progression, increased retention and recommendation to friends and colleagues leading to sustainable workforce and improved clinical care |
| Poor performance and low level of wellbeing in the initial stages at the workplace among new entrant IMGs | Structured intervention during the orientation period including a good learning environment, adequate mentoring and pastoral support will help to overcome these problems | Improvement in the participation, wellbeing, self-efficacy and learning of these doctors |
| Risks to patient safety due to poor knowledge of contextual factors including policies and procedures by the new entrant IMGs | Providing clear communication of curriculum competencies along with peer support and clinical supervision to these doctors | Improvement in the awareness among these doctors of the contextual factors contributing to the provision of high-quality patient care |
| Lack of standardized communication channels between JDs, their supervisors and the trust management | Developing a standardized communication strategy will facilitate effective communication of critical success factors | The stakeholders perceive that their voice is being heard and they are valued resulting in their increased satisfaction |
| Lack of criteria for assessment and unrealistic expectations from new entrant IMGs during their orientation | Tailoring the intervention based on requirements of JDs, their supervisors and managers of the trust | Clarity of roles and requirements and better participation of the stakeholders |
| Lack of exposure during on calls including out of hours work in the initial stages of new entrant IMGs | Increased exposure of IMGs to the acute unselected medical take and out of hours ward cover in a supernumerary role during orientation | Improvement in the self-efficacy of IMGs particularly in dealing with out of hours acute medical emergencies |
| Lack of objective assessments and formal feedback about the performance of new entrant IMGs during initial stages from colleagues | Formal assessment of the performance of new entrant IMGs by medical registrars during supernumerary on calls | Formal assessments provide quality assurance to the programme resulting in increased satisfaction amongst stakeholders about high quality of patient care |

**Table 5:**
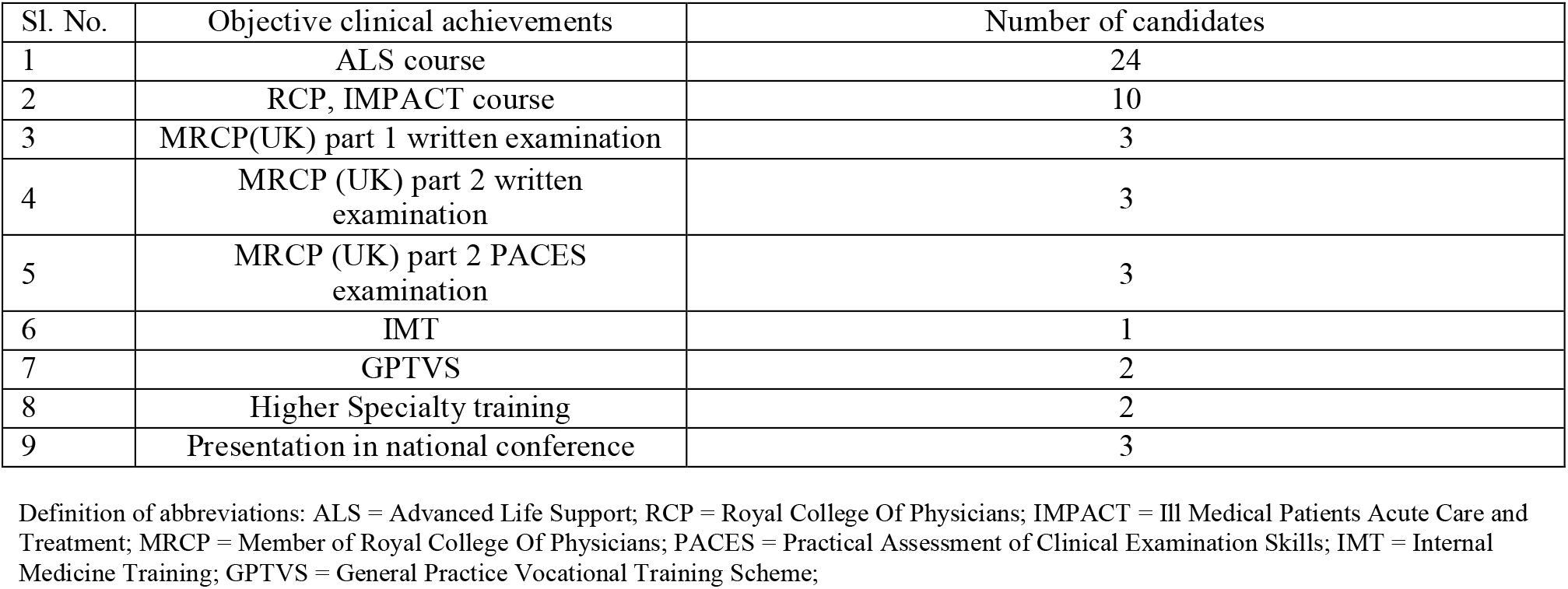
Achievements of new entrant IMGs during employment with the Trust.

| Sl. No. | Objective clinical achievements | Number of candidates |
| --- | --- | --- |
| 1 | ALS course | 24 |
| 2 | RCP, IMPACT course | 10 |
| 3 | MRCP(UK) part 1 written examination | 3 |
| 4 | MRCP (UK) part 2 written examination | 3 |
| 5 | MRCP (UK) part 2 PACES examination | 3 |
| 6 | IMT | 1 |
| 7 | GPTVS | 2 |
| 8 | Higher Specialty training | 2 |
| 9 | Presentation in national conference | 3 |

Definition of abbreviations: ALS = Advanced Life Support; RCP = Royal College Of Physicians; IMPACT = Ill Medical Patients Acute Care and Treatment; MRCP = Member of Royal College Of Physicians; PACES = Practical Assessment of Clinical Examination Skills; IMT = Internal Medicine Training; GPTVS = General Practice Vocational Training Scheme;

## Discussion

To our knowledge, this is the first study to explore the perceptions of new entrant IMGs on the NHS and their peers following a structured orientation programme. Overall perception was positive, corresponding to a stable, well-trained resident doctor workforce which could sufficiently provide assurance of quality of care for patients.

The results, based on realist evaluation, suggest that the COP elements (i.e., Mechanisms) were perceived by the IMGs as effective. Moreover, the IMGs perceived the workplace (i.e., Context) as a supportive learning environment where necessary resources were available and the workload manageable. Regarding Outcomes, IMGs were satisfied with their experiences, were confident to be able to start independent work upon COP completion, remained committed to their employing organization, and would recommend the COP to future colleagues. Overall reliability of the survey was good. This might translate to the current survey being used for a COP adapted to other healthcare organizations nationwide. Most of the IMGs partaking in the COP felt that it should be made mandatory for all future new entrant IMGs. Although there were some suggestions for improvement, the majority stated that those changes had already been implemented by the organizers.

A small minority of IMGs had raised concerns about bullying and harassment in the workplace during their COP. This information was conveyed informally by the IMGs to their mentors who immediately contacted the senior management team of the medicine division who in turn swiftly acted on the concerns and prevented the need for any further escalation. In some instances, IMGs were moved to other departments or to another hospital within the same Trust. The formal peer feedback from the middle grade doctors supervising the new entrant IMGs during the COP was complementary. There were some suggestions for developmental improvement which was wholly constructive in nature and intended for benefit of individual IMGs.

Many of the IMG resident doctors have reached objective professional and academic achievements during their employment with the Trust. Although these achievements may not be comparable to local trainees, most IMGs being in their first year of working in the NHS have demonstrated drive and ambition to develop and deliver on their learning goals. Their achievements are also a reflection of the support provided to them by their supervisors.

In a recent report we have shown that during the intervention (recruitment of IMGs and COP) period between the academic year 2017/2018 and 2018/19, the resident doctor post occupancy rate has risen sustainably, there was a reduction in locum agency spend, formal complaints, exception reporting by trainee residents, and length of hospital stay for inpatients in the medicine division. These factors have led to the study Trust (which includes the medicine division) being rated ‘Good’ (from ‘Requires Improvement’) during the academic year 2018/19, for the first time since its inception by the Care Quality Commission.^8^

The COP has evolved significantly over the past 2 years, and we have continued to refine, optimize and improve the efficiency of the programme. In addition, feedback from participants during the evolution of the programme has enabled the development of a booklet detailing the structure, curriculum, assessment targets and tools provided to the new entrant IMGs and their supervisors at the start of the COP. User-friendly Web-based access to the contents and goals of the programme is under development.

## Limitations

The main limitation of the study was that we could not compare IMG feedback post-intervention with baseline expectations prior to commencement of the programme. This is because nearly half of the resident doctors who were recruited and underwent the COP had also undergone our Trusts’ honorary clinical attachment programme. Therefore, having a baseline survey would have resulted in lack of consistency in the feedback between doctors who had undergone the local attachment programme versus doctors who had not. Despite this, the reflections of the IMGs captured in this study are nonetheless insightful.

## Conclusion

This report confirms that resource investment in COP results in a high degree of satisfaction, workforce retention, and clinical performance of new entrant IMGs, thereby providing direct benefits to quality of patient care. We recommend larger scale implementation of this programme as an acceptable and objective model to strengthen clinical assurance and clinical governance surrounding IMG recruitment.

## Data Availability

All data produced in the present study are available upon reasonable request to the authors.

## Acknowledgements

The authors would like to acknowledge Dr. Alapan Bandyopadhyay, MBBS, DipCMH, Resident at the Department of Community Medicine of North Bengal Medical College and Hospital, India for his help in copy-editing the current article.

